# Using Natural Language Processing to Classify Serious Illness Communication with Oncology Patients

**DOI:** 10.1101/2021.08.20.21262082

**Authors:** Anahita Davoudi, Hegler Tissot, Abigail Doucette, Peter E. Gabriel, Ravi Parikh, Danielle L. Mowery, Stephen Miranda

**Author notes:** shared co-senior authorship (Danielle L. Mowery, Stephen Miranda).

## Abstract

One core measure of healthcare quality set forth by the Institute of Medicine is whether care decisions match patient goals. High-quality “serious illness communication” about patient goals and prognosis is required to support patient-centered decision-making, however current methods are not sensitive enough to measure the quality of this communication or determine whether care delivered matches patient priorities. Natural language processing (NLP) offers an efficient method for identification and evaluation of documented serious illness communication, which could serve as the basis for future quality metrics in oncology and other forms of serious illness. In this study, we trained NLP algorithms to identify and characterize serious illness communication with oncology patients.

## 1 Introduction

For patients with cancer to receive care that aligns with their values, their clinicians must effectively explore their care preferences. Documentation of patient-specific goals and prognostic information earlier in the illness trajectory is critical for assessment of shared decision-making, goal-concordance, and healthcare utilization in oncology. High-quality serious illness communication (SIC) can enhance quality of life and goal-concordant care, ^1,2,3^ while inadequate SIC is associated with greater psychosocial distress and aggressive end-of-life care that may be incongruent with patient preferences. ^4,5,6^ There is consensus that SIC documentation itself is a core quality measure that supports goal-concordance and therefore must be evaluated. ^7,8,9^ However, it is well-documented that traditional forms of SIC documentation, including advance directives, are under-utilized and inconsistently applied, making it difficult to track SIC across inpatient and outpatient settings. ^10,11,12^ High-quality SIC in oncology includes discussion of patient goals, prognosis, code status, and advance care planning. ^13^ Routine assessment of documentation on these four topics is difficult because this information often exists as free-text in the electronic health record (EHR), which requires time-intensive, manual chart review to identify and abstract.

### 1.1 Natural Language Processing

Natural language processing (NLP) can offer an efficient, accurate alternative for identification of SIC in the EHR ^14,15^, and has been used to identify care-planning discussions and palliative care delivery. ^16,17,18^ Despite early progress, more sophisticated approaches are needed to classify and evaluate SIC documentation. At this time, NLP approaches for identification of SIC predominantly rely on keywords derived from chart review. Such lexical approaches lend themselves well to identification of specific care-planning metrics, such as documentation of code status (e.g. “full code”, “do not resuscitate”) and discussions about hospice (e.g. “comfort measures only”). However, these algorithms are limited in their ability to capture nuanced documentation about patient priorities and prognostic communication, which does not always rely on representative keywords, is less prevalent in the EHR, and is highly variable from clinician to clinician, limiting identification of this documentation at scale.

Machine learning approaches that expand beyond keywords may support more accurate and automatic identification of these two critical SIC domains. In this study, we sought to leverage weakly-labeled EHR data from oncology patients to develop and validate an NLP algorithm that automatically identifies and classifies SIC documentation about prognosis and goals.

## 2 Methods

This study was approved by the University of Pennsylvania Institutional Review Board, protocol #842930. We first collected a weakly annotated dataset of free-text entries containing SIC documentation, and then trained several machine learning algorithms to automatically classify SIC documentation by domain and subdomain. Finally, we characterized the features associated with each SIC subdomain.

### 2.1 Dataset and Schema

In 2018, the University of Pennsylvania Abramson Cancer Center implemented the Serious Illness Care Program (SICP) developed by Ariadne Labs, a communication intervention designed to enhance timing, frequency, and quality of shared patient-provider decision-making in oncology. ^19,20^ Oncology clinicians are encouraged to document SIC using an EHR module, which generates a semi-structured “Serious Illness Conversation” note with subheadings by SIC domain. Prior to this implementation, all clinicians at Abramson Cancer Center were instructed to use an “Advanced Care Planning” note template for free-text documentation of SIC. In the new “Serious Illness Conversation” note template, there are nine SIC domains, each with a menu of preset responses to choose from, based on the information acquired from the patient, as well as an optional, free-text comment box to insert free-text that provides more detail. The “Serious Illness Conversation” template outlines nine SIC subdomains, three regarding **prognosis** and six regarding **goals**. The **SIC subdomains** including prompts, the structured *responses* and fictitious, but exemplar free-text statements within the “comments” are listed in **Table 1**.

**Table 1:** Serious Illness Communication Subdomains for Prognosis and Goals.

| Prognosis Domains |  |  |  |
| --- | --- | --- | --- |
| Subdomain | Prompt | Responses | Comment |
| **Prognostic Understanding (PU)** | What is your understanding now of where you are with your illness? | *Overestimates prognosis;*<br>*Accurate understanding of prognosis;*<br>*Underestimates prognosis;*<br>*No understanding of prognosis;* | “He knows he only has weeks to live.” |
| **Information Preferences (IP)** | How much information about what is likely to be ahead with your illness would you like from me? | *Patient wants to be fully informed;*<br>*Patient wants to be informed of big picture, but not details;*<br>*Patient wants some information, but no “bad news”;*<br>*Patient prefers information to be shared with \*\*\** | “She prefers weekly prognosis updates.” |
| **Prognostic Communication (PC)** | Information shared with patient about prognosis | *Uncertain prognosis;*<br>*Possibility of getting sick quickly;*<br>*Limited time, may be as short as \*\*\*;*<br>*May never get stronger or regain function* | “He had questions about prognosis.” |
| Goal Domains |  |  |  |
| Subdomain | Prompt | Responses | Comment |
| **Main Goals (MG)** | If your health situation worsens, what are your most important goals? | *Live as long as possible;*<br>*Pursue every available treatment;*<br>*Avoid hospitalizations/maximize time at home;*<br>*Not be a burden/maintain independence;*<br>*Be physically comfortable;*<br>*Be mentally aware;*<br>*Spent time with family* | “The patient wants to live to see his daughter’s wedding.” |
| **Fears/Worries (FW)** | What are your biggest fears and worries about the future with your health? | *Pain or other symptoms;*<br>*Loss of control or dignity;*<br>*Burdening others;*<br>*Family concerns;*<br>*Financial concerns* | “He worries about becoming dependent.” |
| **Strengths (ST)** | What gives you strength as you think about the future with your illness? | *Friends/family;*<br>*Faith/spirituality;*<br>*Prior experience with adversity* | “Support of family and friends.” |
| **Critical Abilities (CA)** | What abilities are so critical to your life that you cannot imagine living without them? | *Living independently;*<br>*Being mentally aware;*<br>*Interacting with others;*<br>*Dressing, bathing, toileting;*<br>*Eating and drinking* | “Maintaining ability to interact with others is important.” |
| **Tradeoffs (TO)** | If you become sicker, how much are you willing to go through for the possibility of gaining more time? | *Anything to prolong life incl. life support & ICU care;*<br>*Limited hospitalizations, some testing and treatments;*<br>*No further life-prolonging care* | “She doesn’t want to experience any major side effects unless there is a high likelihood of therapeutic benefit.” |
| **Family/Friends (FF)** | How much does your family know about your priorities and wishes? | *Extensive discussion with family about goals and wishes;*<br>*Some discussion, but incomplete;*<br>*No discussion but plans to address these issues;*<br>*No discussion, wants help talking to family;*<br>*Does not want family informed* | “We talked about how he and his wife might begin to have conversations with their daughters.” |

For this study, we queried the Penn Medicine cancer registry for all patients with stage III or IV cancer who were treated across all Penn-affiliated locations and whose records contained “Serious Illness Conversation” notes within our EPIC Clarity electronic data warehouse. Our cohort consisted of 3563 total patients from which 5,145 notes were identified, containing a total of 8,695 distinct “responses” and “comments”. The dataset was randomly split into 6,964 entries (80%) for training and 1,731 entries (20%) for testing.

### 2.2 Serious Illness Communication Classifier Development and Evaluation

Each entry from our dataset was preprocessed using the spaCy library: removing punctuation, eliminating stopwords, reducing case, and encoding n-grams (n=1-3 words).^†^ We also encoded lexical categories using Empath. ^21^ Empath is an unsupervised tool trained using connotations between words leveraging a neural embedding derived from over 1.8 billion words of modern fiction.^‡^ Empath can be utilized to generate lexical categories and contains over 200 built-in, topical and emotional categories generated from common dependency relationships in ConceptNet ^22^ and Parrot. ^23^ Topical categories include *money, home, work, religion, health, death, etc*. Emotional categories include *sadness, anger, positive emotion, negative emotion, etc*. Terms within both categories were verified using Amazon Mechanical Turk reviewers.

Using the comments from our training dataset, we trained four machine learning algorithms: Logistic Regression, XGBoost, BERT, and Bio+Clinical BERT.

- **Logistic Regression** learns a logit regression model that explains the relationship between the features and the class. Our model uses exhaustive grid search and L1-regularization to optimize performance while reducing the likelihood of over-fitting due to few training examples, many irrelevant features, and a large number of parameters.
- **XGBoost (extreme gradient boosted trees)** is a gradient descent algorithm that learns to predict the residual errors of prior models while minimizing the loss of adding new models before unifying models to make a final class prediction. These boosting models optimize speed and accuracy while reducing the likelihood of overfitting by penalizing trees and applying proportional shrinking of leaf nodes. The booster parameter was set to gblinear.
- **BERT (bidirectional encoder representations from transformers)** are pretrained deep bidirectional representations from unlabeled text fine-tuned using a “masked language model” that combines both left and right contexts. ^24§^ We leveraged the pre-trained BERT model to provide the vector representations of the embedding sets. ^25^ The encoded representation is then passed to a drop out layer (drop rate of 0.5).
- **Bio+Clinical BERT** is a BERT model that leverages pre-trained language representations initialized from BioBERT, a BERT model generated from PubMed article abstracts and PubMed Central article full texts ^26^ and then fine-tuned using a clinical corpus of notes (e.g., discharge summaries, physician notes, nursing notes, radiology reports, etc.) from the Medical Information Mart for Intensive Care (MIMIC version III) dataset. ^27^

Using a data-driven approach, we trained each of the four algorithms as a SIC classifier to classify each comment according to SIC domains of goals or prognosis. As a proof-of-concept, we also trained only the logistic regression algorithm to classify 1 out of 9 possible SIC subdomains.

### 2.3 Serious Illness Communication Subdomain Characterization

For each SIC subdomain (e.g., the **Goals** domain has a subdomain of **Strengths**), we applied chi-square feature selection and selected the most significantly associated features (n-grams and Empath categories with p<0.05) associated to each class and applied a log-10 transform to each feature’s p-value. We visualized the associated features by transformed p-value using WordCloud. We also report and compare the distribution of Empath categories across subdomains.

## 3 Results

In this study, we leveraged weakly-labeled EHR data from oncology patients to develop and validate an NLP algorithm that automatically identifies and classifies SIC documentation about prognosis and goals.

In **Figure 1**, we report the distribution of subdomain across the full corpus. Among all free-text comments, 61.4% belonged to the domain goals and 38.6% belonged to prognosis. For subdomains within goals, we observed proportions ranging from 6.3% Strengths to 13.2% Tradeoffs. For subdomains within prognosis, we observed proportions ranging from 7.4% Information Preferences to 17.1% Prognostic Communication.

**Figure 1:**
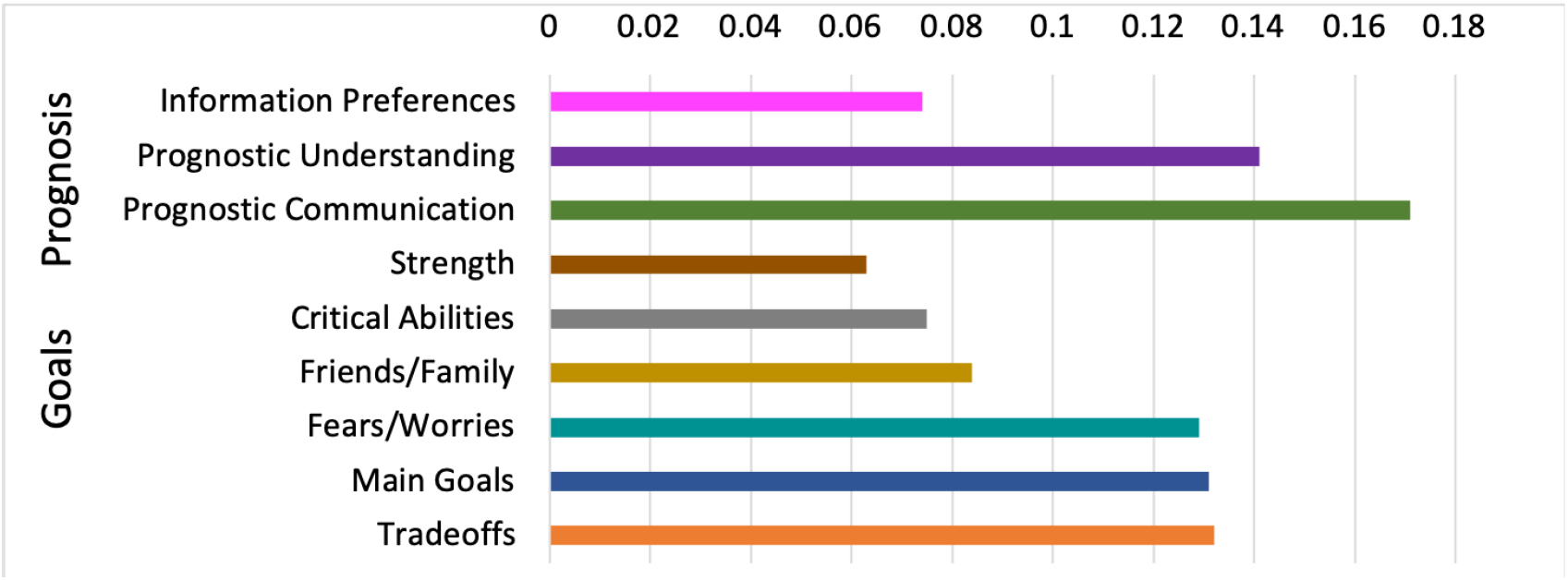
Overall distribution of SIC domains and subdomains.

### 3.1 Serious Illness Communication Classifier Development and Evaluation

In **Table 2**, we report the predictive performance of each machine learning algorithm on the test set. The highest F1-score was achieved by XGBoost for both prognosis (0.86) and goals (0.91). XGBoost achieved the highest precision for prognosis (0.86) and highest recall for goals (0.92). Conversely, Bio+Clinical BERT achieved the highest recall for prognosis (0.86) and highest precision for goals (0.92). In terms of deep learning algorithms, for both prognosis and goals, we observed higher recall (+6 points, +8 points) and precision (+16 points, +2 points) using Bio+Clinical BERT over BERT, respectively.

**Table 2:** SIC classifier performance by SIC domain on the test set.

| **Prognosis** | **Recall** | **Precision** | **F1-score** |
| --- | --- | --- | --- |
| Logistic Regression (baseline) | 0.81 | 0.85 | 0.83 |
| XGBoost | 0.85 | 0.86 | 0.86 |
| BERT | 0.80 | 0.64 | 0.71 |
| Bio+Clinical BERT | 0.86 | 0.80 | 0.83 |
| **Goals** | **Recall** | **Precision** | **F1-score** |
| Logistic Regression (baseline) | 0.91 | 0.89 | 0.90 |
| XGBoost | 0.92 | 0.91 | 0.91 |
| BERT | 0.80 | 0.90 | 0.84 |
| Bio+Clinical BERT | 0.88 | 0.92 | 0.90 |

In **Table 3**, we report the predictive performance of the logistic regression algorithm on the test set for each SIC domain. Among prognosis, the highest F1-score was achieved for Prognostic Understanding (0.61) followed by Prognostic Communication (0.60). Among goals, the highest F1-score was achieved for Critical Abilities (0.71) followed by Strengths (0.65) and Tradeoffs (0.63).

**Table 3:** Logistic Regression SIC classifier performance by SIC subdomain on the test set.

| Prognosis | Recall | Precision | F1-score |
| --- | --- | --- | --- |
| Prognostic Understanding | 0.58 | 0.64 | 0.61 |
| Information Preferences | 0.44 | 0.42 | 0.43 |
| Prognostic Communication | 0.57 | 0.63 | 0.60 |
| Goals | Recall | Precision | F1-score |
| Main Goals | 0.52 | 0.68 | 0.59 |
| Fears/Worries | 0.62 | 0.40 | 0.49 |
| Strengths | 0.75 | 0.58 | 0.65 |
| Critical Abilities | 0.70 | 0.71 | 0.71 |
| Tradeoffs | 0.60 | 0.65 | 0.63 |
| Friends/Family | 0.47 | 0.27 | 0.35 |

### 3.2 Serious Illness Communication Subdomain Characterization

In **Figure 2**, we present the most informative n-grams and Empath categories associated with each SIC subdomain. Features with high associations (low p-values) to a subdomain are larger in the WordCloud. Notable features by prognosis subdomain include: prognostic understanding (*prognosis, understanding, curable, helpful, understands disease, know*), information preferences (*big picture, detail, fully informed*), and prognostic communication (*limited time, short months*). Notable features by goal subdomain include: goals (*spend time, quality, home, live long possible, comfortable*), fears/worries (*fear, concern, loss, dying, suffering*), strength (*strength, friends, catholic, spirituality*), critical abilities (*walking, taking care, reading, independently, self*), tradeoffs (*intubation, dnr, code, life support, would want, measures, considering*), and friends/family (*family, extensive, discussion, wife, daughter, conversation*).

**Figure 2:**
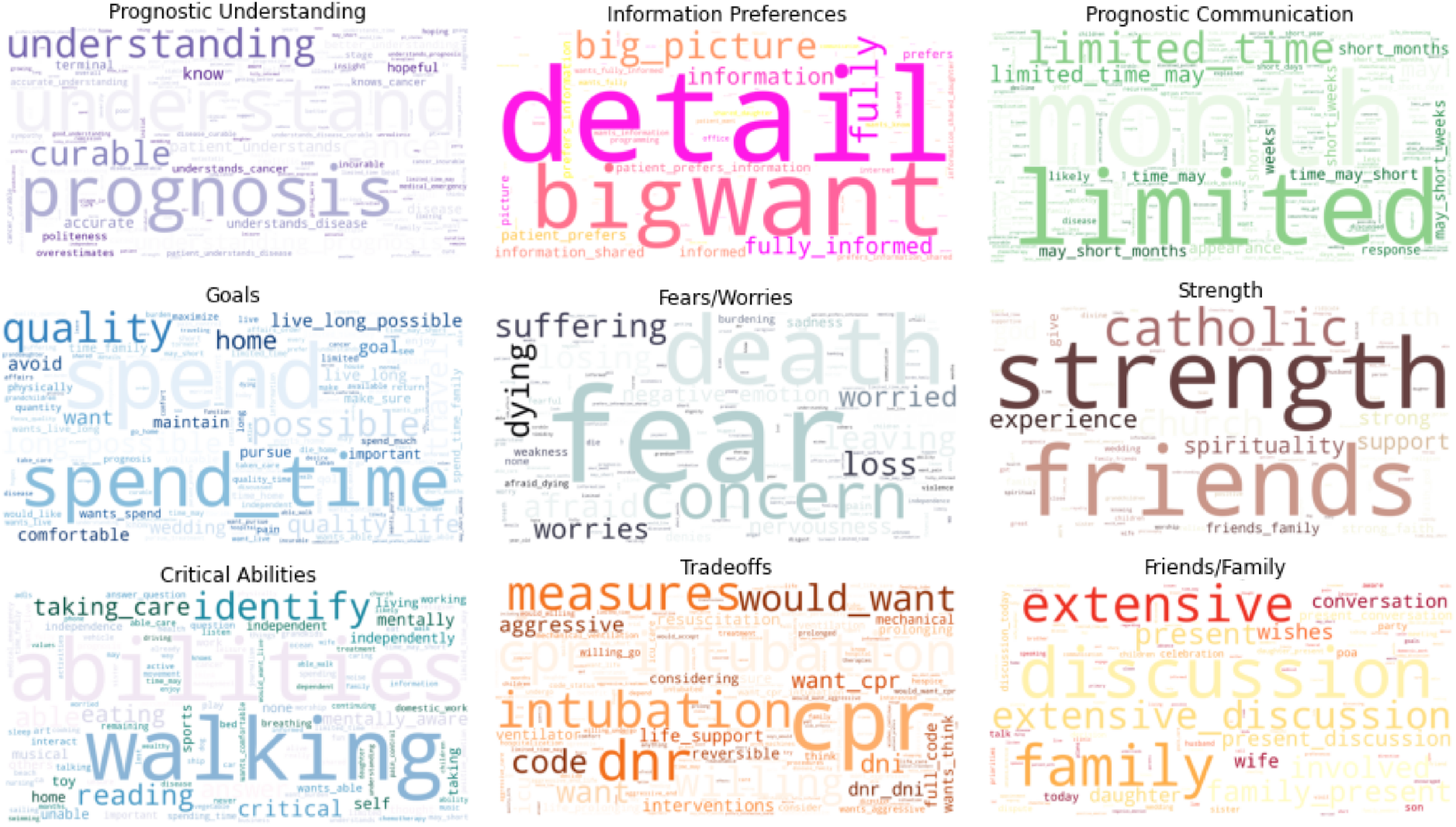
Wordclouds for Prognosis (1st row) and Goal (2nd/3rd row) Subdomains.

In **Figure 3**, we present the frequency distribution of observed Empath categories according to each SIC subdomain. We observed 194 of the more than 200 built-in Empath categories in our full dataset. The most common Empath categories observed across the corpus include: *health, medical emergency, positive emotion, family, children, death, negative emotion*, and *communication*.

**Figure 3:**
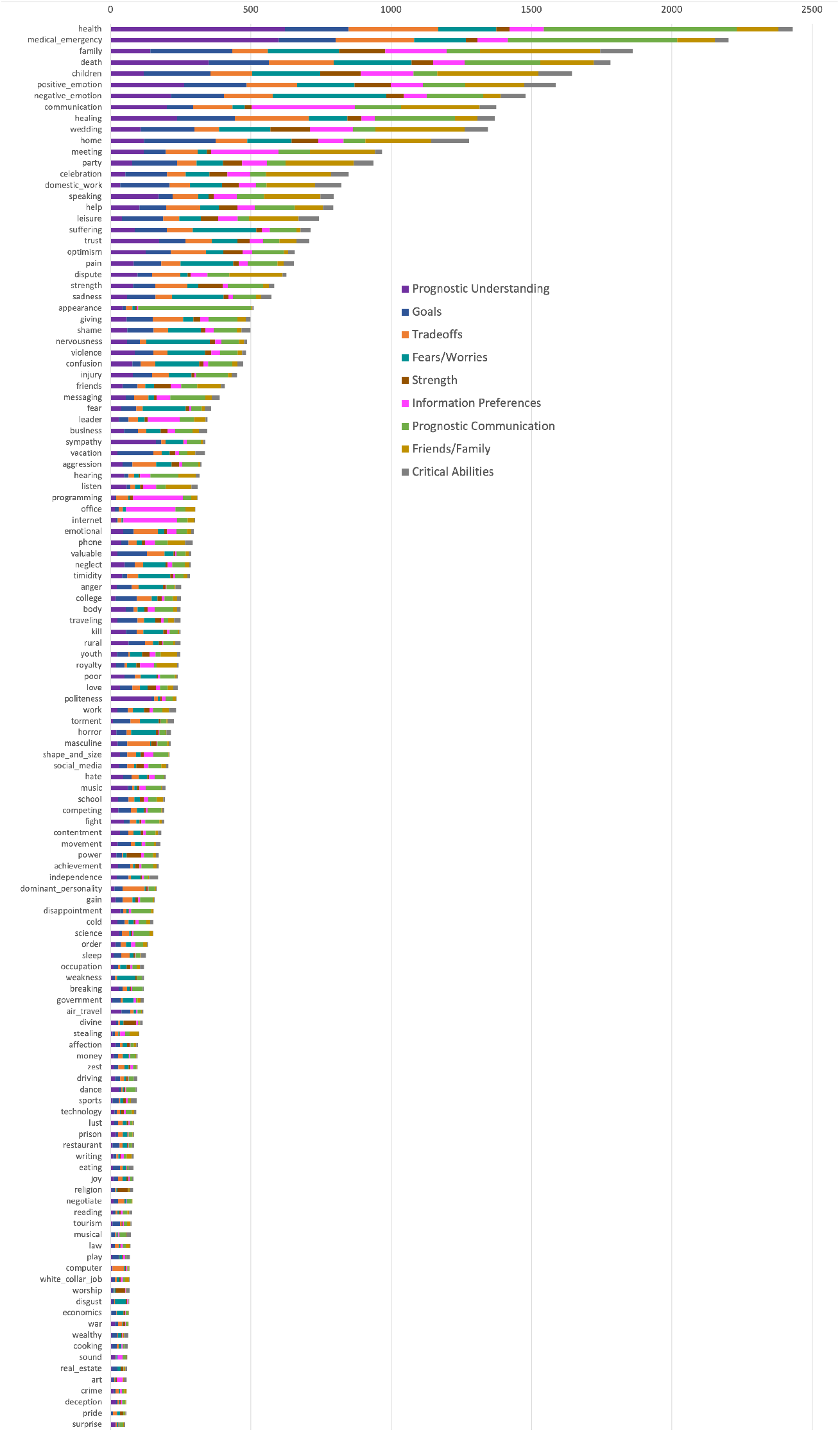
Frequency distributions of subdomains; any Empath category with less than 50 total counts is not shown.

## 4 Discussion

Accurate, reliable, and scalable identification of serious illness communication in the EHR is critical for measuring and improving the quality of oncology care.

### 4.1 Serious Illness Communication Classifier Development and Evaluation

We successfully utilized semi-structured EHR data to develop an NLP algorithm capable of classifying documented entries by SIC domain with high fidelity, identifying text about prognosis (0.86) and goals (0.91). Overall, performance of the classifier across all subdomains ranged from reasonable (0.71) to high (0.91). This study demonstrates promise for identifying SIC—and extracting more complex semantic constructs out of the EHR—without relying on keyword-based approaches. Automated methods for characterizing SIC documentation at scale are limited because clinical notes are variable and often unique to specific clinical situations, which narrow, lexical approaches might fail to anticipate. Here, we leveraged semi-structured data as “weakly labeled” text for classifier training, not only eliminating the need for annotation, but also enhancing the predictive power of the classifier by generating n-grams reflective of diverse lexical categories for training.

The SIC classifier was less effective at discerning individual subdomains within goals and prognosis likely because each subdomain represents overlapping constructs with shared terminology. The “Serious Illness Conversation” template was designed as a communication aid for clinicians to elicit patient values and support prognostic communication, so it is likely that individual subdomains are interrelated for the same patient. While distinguishing between subdomains may be less critical for clinicians using the template at the point of care, enhancing discrimination within each domain would improve classifier performance in free-text clinical notes going forward. It is possible that clinicians inadvertently documented information under the wrong subdomains, which would confound the classifier’s ability to distinguish between them. Notably, classifier performance identified goals better than prognosis, despite a broader range of subdomains, although this may be because the majority of documentation (61.4%) is about goals (**Figure 1**).

The next phase of this research will involve testing and validation of the algorithm’s ability to identify and classify SIC among undifferentiated clinical notes containing unstructured free-text. During this process, further work will be needed to explore why more supervised ML methods (e.g. logistic regression, XGBoost) outperformed deep learning algorithms in this study. Many of the comments and responses used for training and testing consisted of telegraphic phrases, so it may be that deep learning approaches will be more successful in further testing on longer free-text entries, where more contextual features are present. In fact, for both the prognosis and goals domains, we observed higher recall and precision using Bio+Clinical BERT over BERT, respectively, supporting the hypothesis superior performance can be achieved in part through the use of pre-trained models based on clinical documentation.

### 4.2 Serious Illness Communication Subdomain Characterization

Analysis of the most predictive features for each subdomain demonstrates that these features conceptually map very closely to the theme of each subdomain (**Figure 2**) while reflecting a broad range of etymologic categories (**Figure 3**), illustrating the utility of incorporating lexical terms and semantic grouping into the classifier training process. For instance, features associated with documentation about prognosis captured non-specific (*terminal, curable, incurable*) and time-based prognostication (*limited time, short months, short weeks*); the degree of prognostic understanding (*overestimates, accurate, know, good understanding, understands cancer*); how this information was communicated (*office, internet/email*) and to what extent (*detailed, big picture, fully informed*).

Similarly, subdomains within goals, features describe specific wishes or priorities (*wedding, quality time*) and even place of final rest (*home, die house*). Both negative and positive sentiments were reflected. For example, fears/worries contain features of negative emotion (*worried, afraid, suffering, weakness, fearful, nervousness, concern, sadness*); strengths contain features of positive emotion (*comfortable, support, strong*). Sources of strength include one’s faith (*catholic, spirituality, divine*) and support system (*children, friends family*). Critical abilities highlight activities of leisure (*sports, walking, play, art, driving, reading, working*) and daily living (*living, breathing, eating*) as well as terms related to autonomy (*self, independent, dependence*). To achieve these goals and maintain critical abilities, preferences for life-sustaining treatments were also captured, including code status (*intubation, cpr, dnr, life support, resuscitation, ventilation, full code*). Both prognosis and goals were often shared with individuals representing family (*wife, husband, son, daughter, sister, son*) and those in decision-making roles (*poa, power of attorney*).

### 4.3 Clinical Applications

If further validated, the clinical implications of this SIC classifier are compelling. While documentation about goals of care and prognostic communication are known process measures of high-quality palliative care delivery ^28^, SIC is poorly captured by administrative claims data, and manual review of individual patient records is laborious and impractical at the population level—yet quality measurement in palliative care is still highly dependent on these two methodologies. ^12,29,30,31,32^ A validated SIC classifier would offer a powerful tool for more useful quality metrics in oncology, either by evaluating communication quality or developing personalized measures of goal-concordance. ^7,33^ Reliably tracking patient goals would provide useful context for assessing appropriateness of healthcare utilization, and characterizing narrative arcs in the disease trajectory could help frame quality improvement initiatives and psychosocial interventions during serious illness. ^34^ In healthcare operations, explainable AI for logistic regression or XGBoost could even be used to inform clinician-facing EHR tools at the point of care, perhaps by visualizing positive coefficients or SHAP values across terms and Empath categories.

Although these results are preliminary, the methodology employed here allows for greater real-world applicability than other reports of NLP approaches to SIC identification thus far, which have all been keyword-based. ^15,16,17,18^ Recent applications of these methods have seen success in patient groups drawn from pragmatic trials in oncology, ^35,36^ but due to their lexical basis these efforts have required manual annotation of hundreds of clinical notes, and may be weighted towards inpatient admissions or medical crises requiring treatment decisions. ^35^ Our method may lay the foundation for more nuanced identification of patient-specific priorities and prognostic communication more upstream in the disease trajectory, which would have significant utility across a wide array of clinical contexts.

### 4.4 Limitations and Future Work

This study has notable limitations. The SIC classifier was trained using semi-structured EPIC modules, which limits the replicability of this work in other settings where source text enriched with SIC may be lacking. Moreover, most SIC documentation in oncology exists within free-text clinical notes, requiring discrimination between relevant and irrelevant text; performance may suffer in population-level datasets where SIC represents a minority of clinical documentation. In the next phase of this research, we will leverage “Advance Care Planning” notes obtained from our EHR to enhance classifier training for application to free-text clinical notes. This classifier is based on documentation from a limited number of oncology clinicians at one institution requiring further study in larger, more diverse populations to assess generalizability. In the future, we aim to better understand how patient preferences evolve over time, as well as any similarities or differences in SIC across gender, race, ethnicity, and culture. ^37^

## Conclusion

Here we describe a novel application of NLP for classifying SIC documentation in oncology. If further validated, such an algorithm can retrieve and evaluate SIC documentation in routine clinical practice as a quality metric ^38^ to assess key clinical and systems priorities in oncology.

## Data Availability

The dataset is not available.

## Acknowledgements

We extend our gratitude to the University of Pennsylvania for partially supporting this important research through Dr. Mowery’s start-up funding.

## Footnotes

† https://spacy.io/universe

‡ https://github.com/Ejhfast/empath-client

§ https://github.com/google-research/bert

